# Balancing Accuracy and Actionability: An Assessment of Minimal- Input Wastewater Models for COVID-19 Prediction

**DOI:** 10.1101/2025.07.03.25330828

**Authors:** L. M. Lariscy, M. E. Lott, A. Handel, A. M. Foley, C. Melendez-Declet, L. Metsker, E. K. Lipp

## Abstract

Wastewater-based epidemiology (WBE) is a valuable tool for surveillance of infectious diseases like COVID-19, yet balancing prediction accuracy with practical deployment remains challenging, particularly in low-resource, low-prevalence, and early warning contexts. We analyzed a two-and-a-half-year dataset from Athens-Clarke County, Georgia (USA) to compare approaches for predicting COVID-19 cases from SARS-CoV-2 wastewater data. We evaluated the effects of extraction replicates and use of quantitative versus detection frequency data on model performance using random forest and linear models. Results show that two extraction replicates generally suffice for reliable prediction, supporting efficient resource use. Combined viral load and detection frequency produced the strongest models, but detection frequency alone predicted new COVID-19 cases more accurately than viral load alone and was less sensitive to RT-qPCR instrumentation changes, making it a practical alternative when quantification is unreliable/infeasible. Linear models predicted new cases more accurately than random forest models, offering a resource-efficient option for monitoring wastewater trends. Following declines in clinical testing in spring 2022, wastewater-based models estimated substantially higher case counts than were reported, underscoring WBE’s role for ongoing surveillance of COVID-19 and other infectious diseases. These findings provide practical guidance for optimizing WBE implementation, particularly where early warning and resource constraints are significant factors.

**HIGHLIGHTS:**

- Analysis of SARS-CoV-2 in wastewater revealed that presence/absence rates in wastewater predicted new COVID-19 cases better than viral quantity alone
- Two extraction replicates was sufficient for acceptable prediction accuracy
- Linear regression models more accurately predicted new cases than random forest
- After clinical testing efforts diminished, wastewater models predicted higher case counts than were reported

**GRAPHICAL ABSTRACT:** 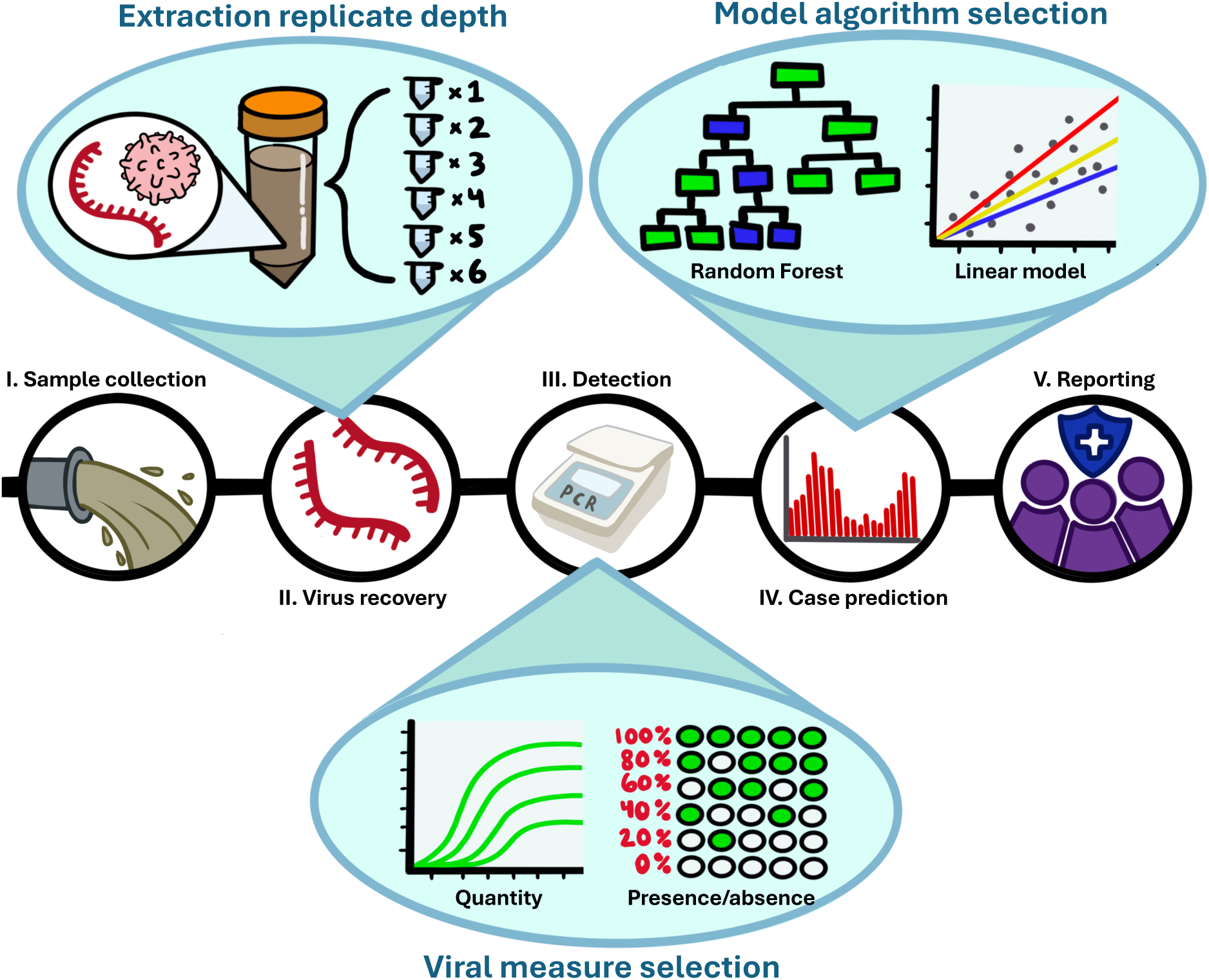

## INTRODUCTION

Wastewater has been used as a tool to track pathogens and inform public health interventions since the 1940s (Moore, 1951), but its potential for tracking infectious disease trends on a wide scale was not fully realized until the emergence of severe acute respiratory syndrome coronavirus 2 (SARS-CoV-2) and subsequent COVID-19 Pandemic in 2020 (Bivins et al., 2020). Early in the pandemic, researchers turned to wastewater surveillance to fill gaps left by traditional surveillance (Ahmed et al., 2020; Haramoto et al., 2020; Randazzo et al., 2020; Sherchan et al., 2020). Infected individuals shed SARS-CoV-2 (Wang et al., 2020; Wu et al., 2020), which can enter sewage streams, and its RNA can be detected downstream through molecular techniques (e.g., RT-quantitative PCR) (Kitajima et al., 2020), a strategy which has been implemented at thousands of sites worldwide (Naughton et al., 2023). In many cases, wastewater surveillance systems can pre-emptively indicate local spikes in cases (Medema et al., 2020; Wu et al., 2022), which can facilitate preparation in healthcare systems and inform public health strategy (Keshaviah et al., 2022). Overall, wastewater-based surveillance is a valuable tool for monitoring community health that overcomes limitations of traditional surveillance (i.e., cost effective, anonymous, not biased by healthcare-seeking behavior) and its utility extends well beyond COVID-19 (Alex-Sanders et al., 2023; Boehm et al., 2024; Stalder et al., 2019; Zhang et al., 2023).

Most SARS-CoV-2 wastewater surveillance programs rely on quantitative PCR technology (i.e., reverse transcription (RT) q-PCR, RT digital droplet PCR) to quantify gene targets within a given sample. While increased viral signals in wastewater can generally indicate increased COVID-19 incidence, this relationship is influenced by a complex web of factors, which complicate analysis and interpretation of results. Several studies have emphasized the importance of these factors for making accurate estimates of community infection rates from wastewater viral detection. These include environmental characteristics (e.g., inflow rate, physiochemical properties) (Li et al., 2023; Schussman & McLellan, 2022), population characteristics (e.g., population size, fecal shedding rate) (Wilder et al., 2021; Wu et al., 2021), and laboratory procedures (e.g., sample handling, extraction replicates) (Ahmed et al., 2022; Weidhaas et al., 2021). While accounting for these factors may contribute to more accurate estimates, the inclusion of a growing range of parameters may increase reporting delays, which can detract from wastewater’s early warning capabilities (Bibby et al., 2021). Additional sample processing and metadata collection may also not be feasible in low-resource settings. To date, the field lacks a critical evaluation of how the pursuit of ever-increasing accuracy may limit the practical utility of wastewater as an accessible, early disease warning system. There is a need to define a set of minimal criteria needed for wastewater-based models to track the status of pathogens such as SARS-CoV-2 within a community; this is particularly important in resource limited settings or where targeted clinical testing may need to be rapidly implemented. Given the current gaps in literature, work is needed to explore the trade-offs of simplified surveillance approaches that strike the right balance between rapid and easy to implement methods, and suitably accurate results.

While most work to date has focused on quantitative indicators of viral presence and abundance in wastewater, there is no standardized approach, making viral load calculations difficult to compare between groups (Bivins et al., 2021), regardless of methods reporting guidelines (Borchardt et al., 2021; Bustin et al., 2009). These quantitative methods may also be limited, as viral signals are often undetectable or outside the range of quantification (Ahmed et al., 2022; Zhu et al., 2022). However, presence/absence (detection frequency) of SARS-CoV- 2 in wastewater has been shown to adequately reflect COVID-19 incidence (Bivins et al., 2022; Lott et al., 2023; Wang et al., 2025) and may provide a valuable alternative when quantifiable data is unattainable or unreliable (Zhao et al., 2023; Zhu et al., 2022). Despite its potential for use in low-prevalence or low-resource settings, detection frequency remains underutilized in predictive models. Relying solely on quantitative techniques may leave behind informative data that are easier to collect, deploy in low-resource settings, and standardize than quantifiable data like viral load or concentration (Bivins et al., 2022). To address these gaps, this study evaluates how different SARS-CoV-2 wastewater surveillance methodologies impact the accuracy of COVID-19 case predictions. Specifically, we assess the effects of 1) extraction replicate depth, 2) quantitative vs qualitative (presence/absence) viral metrics, and 3) model algorithm complexity, with a focus on identifying approaches that support timely, resource-efficient disease monitoring.

## METHODS

### Setting

Wastewater surveillance was conducted at three water reclamation facilities in Athens-Clarke County, Georgia, United States (Figure 1). Approximately 131,000 total residents are served by these treatment facilities. The largest facility (Plant A) serves about 56,500 residents, including the University of Georgia main campus and the Downtown Athens area, as well as industrial waste sources. The second largest facility (Plant B) serves about 49,000 residents, as well as two major hospitals. The smallest facility (Plant C) serves about 29,000 residents and the catchment zone is primarily residential, but input is also received from pumped septic systems and portable restrooms from neighboring counties (Athens-Clarke County Public Utilities Dept., 2020).

**Figure 1.**
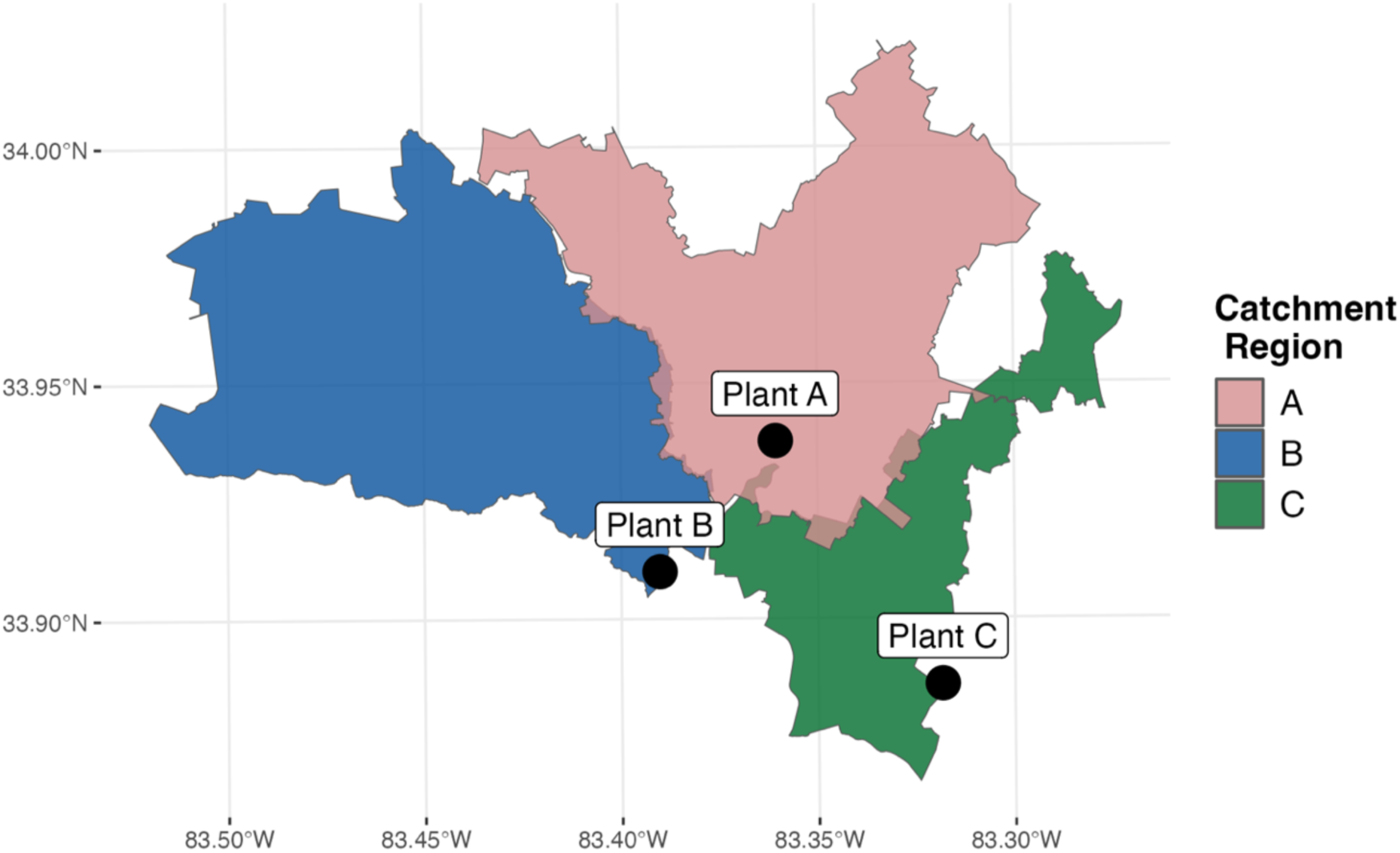
Wastewater catchment regions of each sampled reclamation facility.

### Wastewater sample collection

Sample collection for wastewater surveillance began June 30, 2020, and occurred twice weekly (Mondays, Wednesdays) through January 4, 2023. During this surveillance period, a 24-h time composite raw wastewater influent sample (1L), representative of influent from 6 a.m. the prior day through 6 a.m. of the date of collection, was collected from each of the three WRFs and stored at 4°C until ready for extraction (< 3 h). Wastewater influent flow data, including millions of gallons per day (MGD) and total suspended solids (TSS), was provided by plant operators for each sampling period. Wastewater flow rates were converted from MGD to liters per day prior to downstream analysis.

### Purification of viral RNA

Viral RNA extraction methods are detailed in Lott et al. (2023). In summary, the QiAmp Viral RNA Mini Kit (catalog no. 52906, Qiagen) was used to purify RNA directly from unconcentrated wastewater samples, processing controls, and extraction blanks (molecular-grade water) in 280 µl volumes following manufacturer instructions. For each wastewater sample, up to six replicates were extracted, followed by storage at 4°C prior to RT-qPCR (< 24 h).

### Process controls

Sample process controls were prepared as described in Lott et al. (2023). Briefly, bovine coronavirus (BCoV) carried in CalfGuard was spiked into a subsample of each influent sample (1.0 × 10^4^ gene copies ml^−1^) (Lott et al., 2023), followed by RNA purification (described above) and quantification by RT-qPCR using a BCoV targeted assay (Lott et al., 2023). Process controls were then used to estimate viral recovery from direct extraction and RT-qPCR (Lott et al. 2023).

### Reverse transcription and quantification by qPCR

For samples collected in year 1 (June 30, 2020 – June 30, 2021), methods for two-step RT-qPCR and standard curve calculations are described in Lott et al. (2023). Briefly, each purified RNA sample was converted to cDNA using a random hexamer primer and reverse transcriptase (Lott et al., 2023), followed by real-time quantitative PCR (qPCR) on a StepOne Real-Time PCR system (in triplicate) to determine quantification cycle (Cq) for detection of SARS-CoV-2 N1 and N2 genes (Lott et al., 2023).

For samples collected after June 30, 2021 (year 2+), quantification was conducted using one-step RT-qPCR on the CFX384 Touch Real-Time PCR Detection System (BioRad). For each extraction replicate and gene target (N1 and N2), 5 µl of purified RNA, 5 µl of TaqPath 1-Step RT-qPCR MM (catalog no. A15299, Life Technologies), 1.5 µl of 2019-nCoV (N1/N2) premixed primers and probes (0.5 µM [forward], 0.5 µM [reverse], 0.13 µM [probe], final concentrations) (catalog no. 10006770, Integrated DNA Technologies), and 8.5 µl of molecular biology grade water (catalog no. 46-000-CM, Corning) were combined for a final reaction volume of 20 µl. Reactions were run (in triplicate) for each extraction replicate and gene target under the following conditions: 25 C for 2 min, followed by 40 cycles of 50 C for 15 s, 95 C for 2 min, 95 C for 3 s, and 55 C for 30 s.

### SARS-CoV-2 positive control

Details about the positive control used for both surveillance years can be found in Lott et al. (2023). In short, synthetic RNA fragments containing SARS-CoV-2 N1 and N2 gene sequences were assayed in parallel with samples under the same reaction conditions (for each year respectively) (Lott et al., 2023).

### Standard curves

Details about standards used for both surveillance years can be found in Lott et al. (2023). A SARS-CoV-2 plasmid standard, including both N1 and N2 genes, was linearized and used to generate a standard curve for both N gene targets (Figure S1) (Lott et al., 2023). A BCoV DNA Ultramer standard was used to generate a standard curve for the process controls (Lott et al., 2023). Due to instrumentation change, standard curves for each gene target were generated using both the StepOne system (year 1) and the BioRad CFX system (year 2+) (Figure S1). Standards were assayed under the same reaction conditions as samples, for each year respectively.

### RT-qPCR inhibition control

To assess inhibition, sample RNA was spiked with BCoV standard. The BCoV standard (in molecular biology grade water) was assayed in parallel with spiked samples. Samples were determined to be inhibited if Cq values from the sample spiked with BCoV and the molecular biology grade water spiked BCoV were more than 3.33 cycles apart (Lott et al., 2023) (Figure S3; Figure S4).

### RT-qPCR reporting quality

We address the Environmental Microbiology Minimum Information (EMMI) Guidelines (Borchardt et al., 2021) in Table S14 (Supplementary Material).

### Limit of detection and quantification

The limit of detection (LoD) and limit of quantification (LoQ) were estimated by determining deviations in Cq values from a normal distribution of all data points generated in this study, as described in Lott et al. (2023). The LOD and LOQ was determined for each unique assay (i.e., N1 year 1, N1 year 2+, N2 year 1, and N2 year 2+) (Figure S2; Table S5; Table S6).

### Calculation of sample concentration

For reactions with quantifiable signals (Cq > LoQ), copies per microliter (copies µl^-1^) of each gene target in each reaction (C_RXN_) was determined by transforming the Cq value with the appropriate standard curve equation (Figure S1). For non-quantifiable but detectable reactions above the LoD (LoD < Cq < LoQ), concentration values were assigned as half of the LoQ value (copies µl^-1^). For reactions with no detection or that were below the LoD (Cq < LoD), concentration values were assigned as half of the LoD value (copies µl^-1^). Viral concentration was calculated as copies per liter (copies L^-1^) of influent for each gene target (C_WW_) according to Lott et al. (2023) (year 1) and Equation 1 (year 2+).

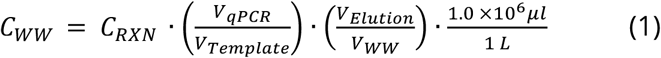

Equation 1 **|** Calculation of copies L^-1^ (C_WW_). Variables accounted for include the initial wastewater input volume, V_WW_ (280 μl), the elution volume, V_Elution_ (60 μl), the volume of template in the qPCR reaction, V_Template_ (5 μl), and the total volume of the qPCR reaction, V_qPCR_ (20 μl).

### Viral load calculation

Due to differences in LoD and LoQ values between year 1 and year 2+, the year 2+ LoD and LoQ were also used for substitutions of year 1 nonquantifiable and non-detection data points (i.e., N1 year 2+ used for N1 year 1). For each sampling date and treatment facility, the wastewater flow rate (liters d^-1^) was used to normalize concentrations and account for sewer flow dilutions, generating the total viral load (VL) of each gene target per wastewater catchment zone per day (Lott et al., 2023). Prior to analysis, average weekly VL per catchment was determined for both N1 and N2 gene targets by taking the geometric mean of each measurement per week (Table S7).

### Detection frequency calculation

Detection frequency (DF) was determined by calculating the number of positive detections per week out of the total number of reactions performed for each catchment zone and gene target (Table S8). This included all sampling events per week (n=2), extraction replicates (n=6), and qPCR technical replicates (n=3) (up to 36 reactions).

### COVID-19 clinical data

Daily reported COVID-19 clinical data for Athens-Clarke County were obtained from the Georgia Department of Public Health (Georgia Department of Public Health, 2024) and used to determine county-wide reported cases per 100,000 for each week in the surveillance period (Table S1). This metric of COVID-19 incidence was used as the outcome variable in this study upon which predictions were made. Data were normalized by taking the base-10 logarithm (log_10_) prior to any model training.

### Random selection of extraction replicates

Prior to any data summarization, extraction replicates corresponding to each original sample were randomly selected from the original dataset at n=1, 2, 3, 4, 5, & 6 to simulate simplified wastewater surveillance processing. At most, six extraction replicates were extracted during surveillance, making n=6 equivalent to the original dataset.

### Data splitting and predictor variable selection

Surveillance data collected prior to the last week of 2021 was selected for model training. This was due to a significant decline in clinical testing efforts after Spring 2022 (Malhotra, 2022) (Figure 2b), which could heavily skew the relationship between wastewater signals and reported cases if used to train models. Data collected in early 2022, prior to testing decline, were used in the test set for model validation.

**Figure 2.**
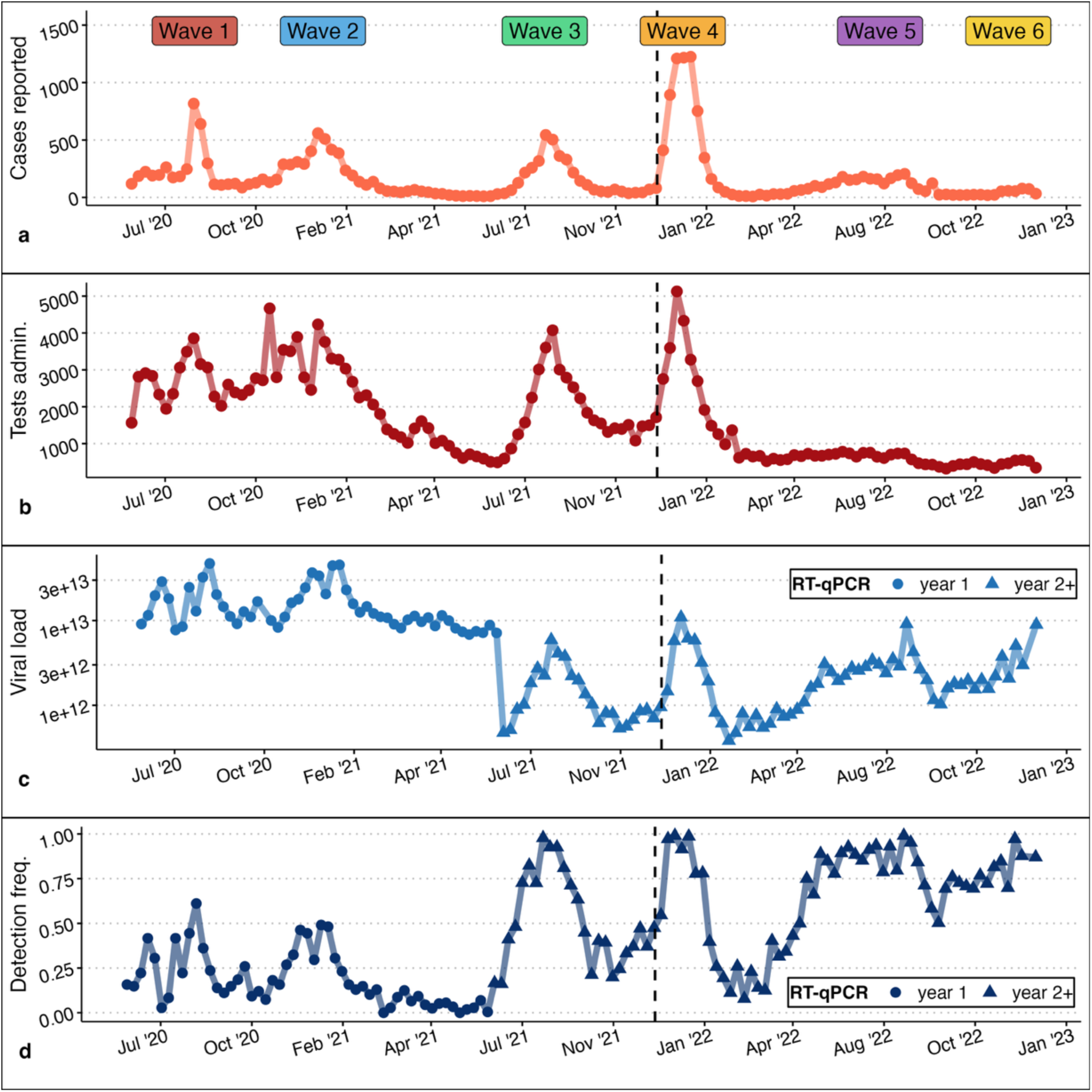
Timeseries of (a) cases reported and (b) tests administered during the surveillance period (per week per 100,000 individuals), as well as (c) average weekly SARS-CoV-2 viral copies in wastewater and (d) total weekly SARS-CoV-2 detection frequency in wastewater. X-axis labels represent the end of each month labeled. Observations to the left of the vertical line were used in model training.

To better understand the utility of wastewater detection frequency as a predictor of COVID-19 incidence compared to a common wastewater viral quantification metric (viral load), three sets of predictor variables were generated for comparison: average weekly wastewater viral load (VL), total weekly wastewater detection frequency (DF), and a combination of these predictors (ALL) (Table 2). For each predictor selection, data from each of the three WRFs and both gene targets were included (Table 1).

**Table 1.** SARS-CoV-2 wastewater variables used to predict COVID-19 cases, including measures of viral presence in wastewater, wastewater catchment zone, and SARS-CoV-2 gene target.

| Variable | Wastewater viral measure | Catchment | Gene target |
| --- | --- | --- | --- |
| WW<sub>1</sub> | log<sub>10</sub>(average weekly wastewater viral load) (VL) | Plant A | N1 |
| WW<sub>2</sub> |  |  | N2 |
| WW<sub>3</sub> |  | Plant B | N1 |
| WW<sub>4</sub> |  |  | N2 |
| WW<sub>5</sub> |  | Plant C | N1 |
| WW<sub>6</sub> |  |  | N2 |
| WW<sub>7</sub> | Total weekly wastewater detection frequency (DF) | Plant A | N1 |
| WW<sub>8</sub> |  |  | N2 |
| WW<sub>9</sub> |  | Plant B | N1 |
| WW<sub>10</sub> |  |  | N2 |
| WW<sub>11</sub> |  | Plant C | N1 |
| WW<sub>12</sub> |  |  | N2 |

**Table 2.** All models tested, including unique combinations of model types, predictor sets, and extraction replicate depths.

| Model type | Predictor set | Sample rep. | Model name |
| --- | --- | --- | --- |
| Linear model (LM) | Viral load (VL) | 1 | LM VL 1 |
|  |  | 2 | LM VL 2 |
|  |  | 3 | LM VL 3 |
|  |  | 4 | LM VL 4 |
|  |  | 5 | LM VL 5 |
|  |  | 6 | LM VL 6 |
|  | Detection frequency (DF) | 1 | LM DF 1 |
|  |  | 2 | LM DF 2 |
|  |  | 3 | LM DF 3 |
|  |  | 4 | LM DF 4 |
|  |  | 5 | LM DF 5 |
|  |  | 6 | LM DF 6 |
|  | Viral load + detection frequency (ALL) | 1 | LM ALL 1 |
|  |  | 2 | LM ALL 2 |
|  |  | 3 | LM ALL 3 |
|  |  | 4 | LM ALL 4 |
|  |  | 5 | LM ALL 5 |
|  |  | 6 | LM ALL 6 |
| Random forest (RF) | Viral load (VL) | 1 | RF VL 1 |
|  |  | 2 | RF VL 2 |
|  |  | 3 | RF VL 3 |
|  |  | 4 | RF VL 4 |
|  |  | 5 | RF VL 5 |
|  |  | 6 | RF VL 6 |
|  | Detection frequency (DF) | 1 | RF DF 1 |
|  |  | 2 | RF DF 2 |
|  |  | 3 | RF DF 3 |
|  |  | 4 | RF DF 4 |
|  |  | 5 | RF DF 5 |
|  |  | 6 | RF DF 6 |
|  | Viral load + detection frequency (ALL) | 1 | RF ALL 1 |
|  |  | 2 | RF ALL 2 |
|  |  | 3 | RF ALL 3 |
|  |  | 4 | RF ALL 4 |
|  |  | 5 | RF ALL 5 |
|  |  | 6 | RF ALL 6 |
| Null Model | Combined (ALL) | 6 | NULL |

To test the impact of replicate depth on prediction capabilities across the three main predictor sets (VL, DF, ALL), extraction replicate subsets (n=1, 2, 3, 4, 5, 6) were used to generate six unique predictor sets for each of the three main predictor sets. This resulted in 18 unique predictor sets (Table 2).

### Correlation analysis

Data generated from wastewater surveillance and clinical reporting were processed as described above and relationships were evaluated using univariate Spearman’s correlations, using all extraction replicates. Correlations were assessed between wastewater-generated SARS-CoV-2 measures (average weekly viral load, and total weekly detection frequency) and clinical-generated COVID-19 measures (weekly reported cases/100,000) for each WRF and gene target.

Correlations were also assessed between the two wastewater measures. Correlations were first examined on model training data and test data (separately) to examine how relationships between wastewater and clinical variables changed when case reporting efforts diminished (Figure S5a; Figure S5b). Following this, correlations were then evaluated on year 1 and year 2+ separately to account for RT-qPCR assay discrepancies and to better examine correlations between wastewater variables (Figure S5c; Figure S5d).

### Model tuning

Linear regression was selected as a ‘simple’ model, representing a straightforward and computationally light application. Random forest was selected as a more ‘complex’ model, a framework which has been previously explored for use in wastewater-based models and has been shown to explain a substantial portion of the variance in observed data (Koureas et al., 2021; Rezaeitavabe et al., 2025), indicating reasonably accurate predictions. Random forest hyperparameters were tuned separately for VL, DF, and VL+DF (ALL) models using all extraction replicates in the training dataset. A tuning grid of 20 automatically generated candidate parameters was utilized alongside 10-fold cross validation to determine optimal hyperparameters based on R^2^ values (Table S9). Finalized models with optimized hyperparameters were combined in workflows with each unique predictor set. Linear regression models using ordinary least squares were also combined in workflows with each unique predictor set (Equations 2 – 4). All finalized models (Table 1) were fit to resamples of the training data.

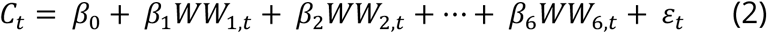

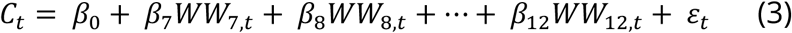

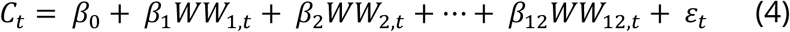

Equations 2 – 4 **|** Linear models for main wastewater predictor sets, including viral load (VL) (2), detection frequency (DF) (3) and VL+DF (ALL) (4), where: *Ct = log_10_(weekly cases per 100,000)*, *β_0_ = y-intercept*, *β_1_, β_2_, … β_12_ = slope coefficients*, *WW_1,t_, WW_2,t_, … WW_12,t_ = wastewater predictor variables* (Table 1), and *ε_t_ = error*.

### Model comparison

To compare each model, 10-fold cross validation with 10 repeats was used to evaluate model performances on the finalized workflows. R-square (R^2^) and root mean square error (RMSE) of observed versus predicted values were calculated for each fold and repeat (100 total) to evaluate model performance (Table S10). Performance metrics were calculated after returning the dependent variable to its original unit. Additionally, null models were generated using the ‘ALL 6’ (all wastewater predictor variables and all extraction replicates) dataset, followed by calculation of average cross-validated performance metrics (Table S10).

### Final evaluation on test data

To compare the predictive capabilities of more complex wastewater models versus simplified approaches, we used models fit with each of the predictor sets in both random forest and linear regression frameworks to make predictions on new data (2022). Based on cross-validated performance results (Table S10), which indicated that more than two replicates did not substantially improve performance, only models with two extraction replicates were selected for further testing, yielding six models (Figure 4).

**Figure 3.**
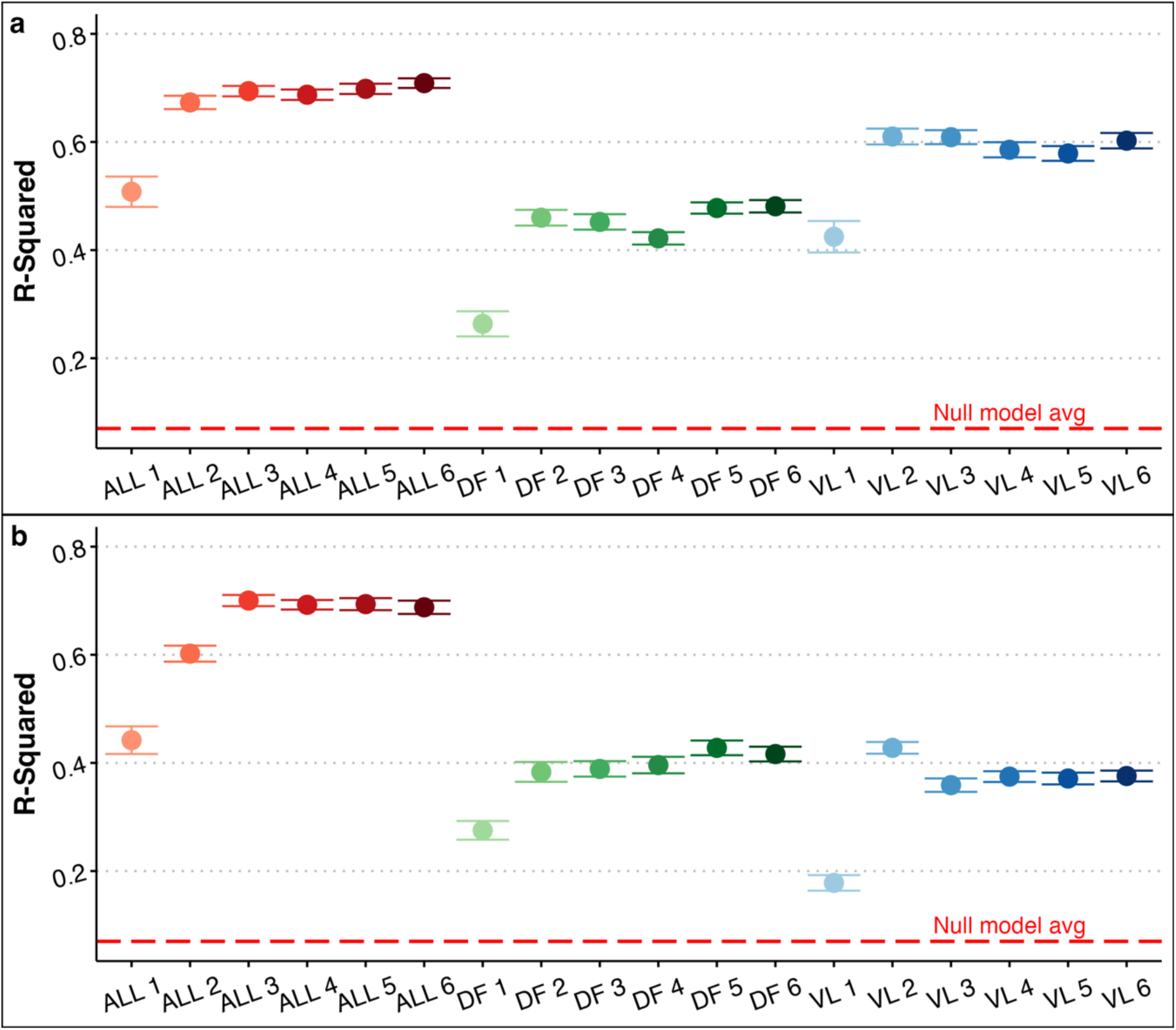
Average cross-validated R^2^ and 95% CI of all predictor sets evaluated in (a) random forest (b) linear models (fit to resamples of model training data). This includes models trained on wastewater viral load (VL), wastewater detection frequency (DF), and combined DF-VL (ALL) predictor sets, as well as randomly selected extraction replicates (1-6).

**Figure 4.**
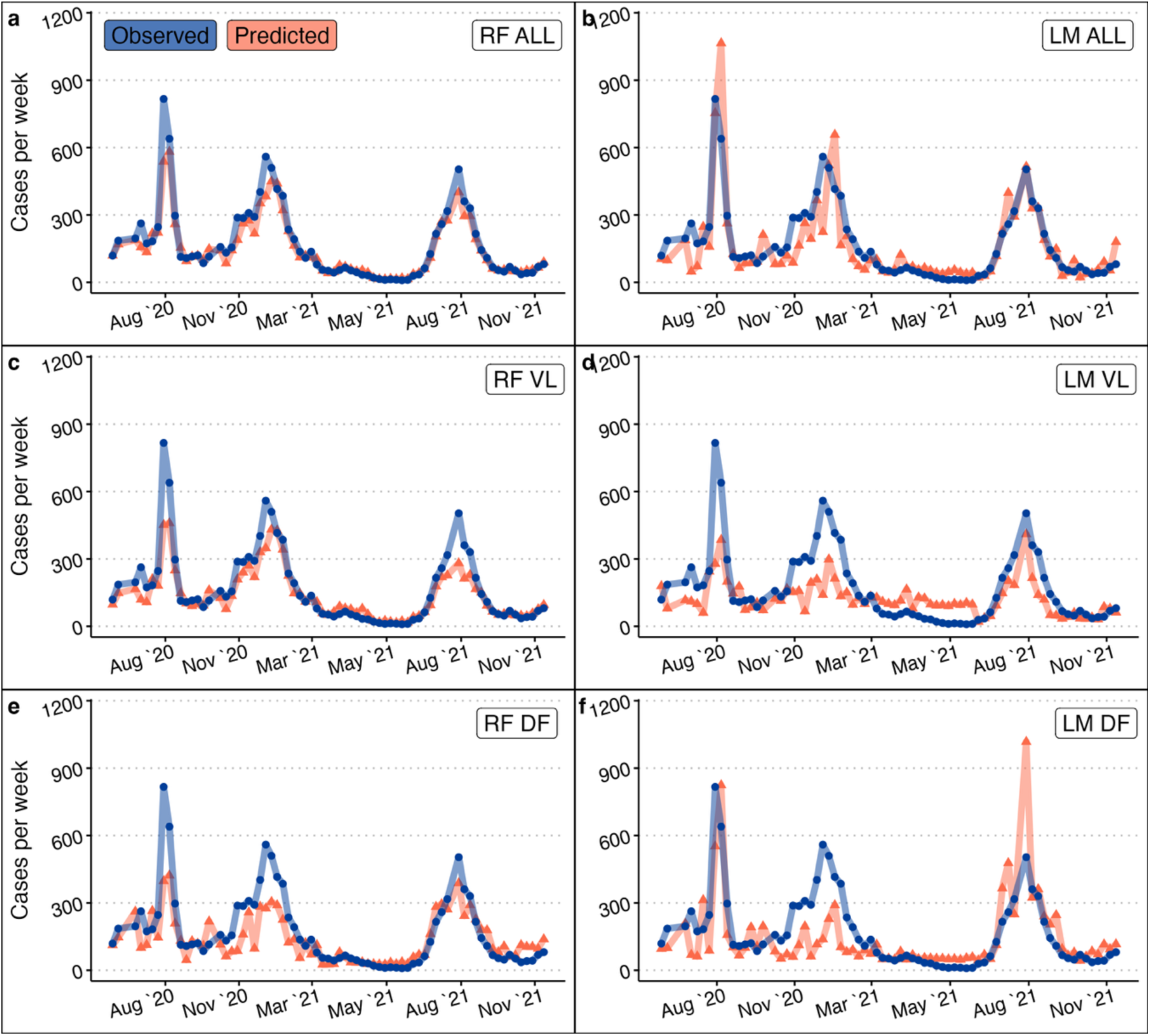
Timeseries of predicted and observed reported cases per week per 100,000 individuals from 2020 to 2021 (model training data). Predictions were made using six models: random forest models with ALL (viral load + detection frequency) (a), VL (viral load) (c), and DF (detection frequency) (e) predictor sets, and linear regression models with ALL (b), VL (d), and DF (f) predictor sets. All models were trained on data generated from two extraction replicates.

### Implementation

All data wrangling and analyses were conducted with R (v.4.4.1) in RStudio (v.2024.09.1+394) using tidyverse (v.2.0.0) (Wickham et al., 2019) and tidymodels (v.1.2.0) (Khun & Wickham, 2020) packages. All data and scripts are available on Zenodo (https://doi.org/10.5281/zenodo.15635445).

## RESULTS

### COVID-19 community prevalence

Throughout the study period (June 30, 2020 – January 4, 2023), a total of 29,869 cases of COVID-19 were reported in Athens-Clarke County, capturing five pandemic waves likely corresponding with the rise of Alpha (B.1.1.7), Beta (B.1.351), Delta (B.1.617.2), Omicron (B.1.1.529), and subsequent Omicron variants (BA.4, BA.5, XBB). The weeks with the highest number of reported cases occurred in early January 2022 (1,576; week of January 16, 2022) and weeks with the lowest reported cases occurred in June 2021 (12; week of June 20, 2021) and March 2022 (10; week of March 20, 2022) (Figure 2a). Weeks with the highest reported cases were also the weeks with the highest number of tests administered (6,599; week of January 2, 2022). While test administration was high during late summer/fall of 2020 (6,007; week of November 15, 2020) and 2021 (5,244; week of August 31, 2021), the 14 lowest testing weeks in the study period occurred in the fall of 2022 (414; week of October 2, 2022), indicating reduced testing (Figure 2b).

### SARS-CoV-2 prevalence in wastewater

A total of 692 wastewater samples were collected over a period of 131 weeks. A maximum of six replicates were extracted from each sample and analyzed for SARS-CoV-2 prevalence using two RT-qPCR targeted assays (N1, N2) across two RT-qPCR platforms (Y1, Y2+), resulting in a total of 6,868 reactions (916 (N1 Y1), 914 (N2 Y1), 2548 (N1 Y2+), 2490 (N2 Y2+)) (Table S3; Table S4). Due to supply chain issues, some extracts (<10%) were stored for >24 hours prior to RT-qPCR. A reaction was considered positive if at least one technical replicate (n = 3) had a Cq value below the Cq LoD. Out of the total reactions, 4,495 (65.4%) were positive (in Year 1: 29.5% (N1), 29.8% (N2); in Year 2+ 86.1% (N1), 70.6% (N2)) (Table S3). Of the total positive reactions, 4,366 (97.1%) were quantifiable (Cq < Cq LoQ) (Year 1 81.1% (N1), 83.5% (N2); Year 2+ 100% (N1), 98.1% (N2)) (Table S3). While ranges in Cq values were similar across all assays (Table S2), detection and quantification limits were notably lower in year 2+ (Figure S2; Table S5; Table S6). Switching from the StepOne system (and two-step assay) to the BioRad CFX system (and one-step assay) decreased limits of detection and quantification (copies L^-1^) in both gene targets, allowing for detection and quantification at lower concentrations.

Weekly average viral load estimates ranged from 1.85 x 10^11^ to 1.33 x 10^14^ N1 copies d^-1^ and 2.99 x 10^11^ to 9.37 x 10^14^ N2 copies d^-1^ (Figure 2c; Table S7).

Positive detection frequency was determined using each extraction replicate (rate of positive reactions) and aggregated per sample. Weekly total detection frequency ranged from 0-100% detection across all treatment facilities and gene targets (Figure 2d; Table S8).

### Correlation analysis

Discrepancies in RT-qPCR methods between year 1 and 2+ made it difficult to assess correlations between variables when data from both years were combined (Figure S5a; Figure S5b). Thus, relationships between variables were assessed in each year separately (Figure S5c; Figure S5d). Across both years, we found the strongest correlations existed between wastewater viral load and detection frequency counterparts (i.e., same catchment and gene target), with stronger correlations on average in Year 2+ (r = 0.95) than in Year 1 (r = 0.88).

This is likely due to improved limits of detection and quantification in Year 2+, allowing for better characterization of wastewater viral presence (Figure S2; Table S5; Table S6).

Conversely, correlations between wastewater variables and reported cases were higher on average (across all sites and gene targets) in Year 1 (r = 0.65) than in Year 2+ (r= 0.52). Although detection and quantification of SARS-CoV-2 in wastewater was improved in Year 2+, this decrease is likely due to diminished clinical reporting and testing during this time (Figure 2a; Figure 2b). Wastewater detection frequency and viral load in Year 1 showed similar correlations on average with case report data (r = 0.66 and 0.64, respectively), but in Year 2+ the association of detection frequency with case reports was stronger (r = 0.55) than correlations of viral load with case data (r = 0.49).

### Model comparison

Across modeling frameworks and feature selections, R^2^ (when fit to training data resamples) tended to increase as replicate depth increased; however, this increase was most notable moving from a single replicate to two replicates, with mean R^2^ values increasing by 0.10 to 0.25 (Figure 3; Table S10), depending on predictor variable-model framework combination. This scale of increase was not observed between other replicate depths (2-6) (Figure 3; Table S10). These results suggest that the greatest gains in model performance from increased replicates occur with the inclusion of a second replicate, after which improvements plateau.

Random forest (RF) and linear models (LM) with both wastewater viral load and detection frequency as predictors (Combined (ALL)) performed best overall. Combined models performed best when evaluated on training data resamples (CV Mean R^2^: 0.66 (RF ALL), 0.64 (LM ALL)) (Figure 3; Figure 4; Figure S6; Table S10; Table S11), but detection frequency (DF) models performed nearly as well when evaluated on new data (Test R^2^: 0.86 (RF ALL), 0.74 (RF DF), 0.79 (LM ALL), 0.8 (LM DF)) (Figure 5; Figure 6; Table S12). Viral load models performed similarly to detection frequency models when fit to training data resamples (CV Mean R^2^: 0.43 (RF DF), 0.57 (RF VL), 0.38 (LM DF), 0.35 (LM VL)) (Figure 3; Figure 4; Figure S6; Table S10; Table S11), but performed worse when predicting new data (Test R^2^: 0.65 (RF VL), 0.57 (LM VL)) (Figure 5; Figure 6; Table S12). All models tested outperformed the null model (CV Mean R^2^: 0.07) (Figure 3; Figure S6; Table S10), which simply predicted the mean of the outcome variable for all observations. In short, models incorporating both wastewater measures demonstrated the best performance, but detection frequency alone offered a simplified, generalizable alternative.

**Figure 5.**
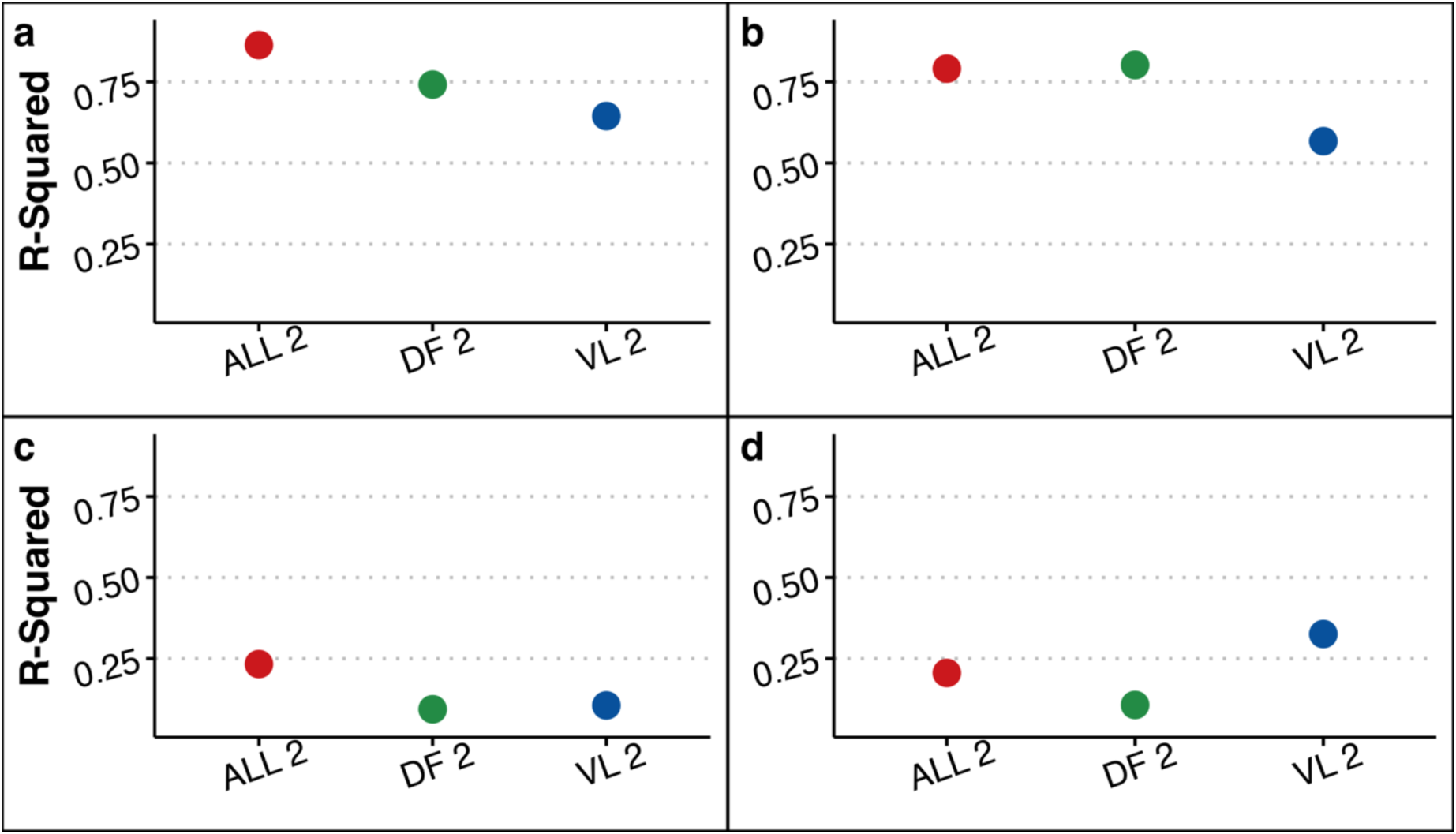
R^2^ of all predictor sets with two extraction replicates evaluated in random forest (a, c) and linear models (b, d) (fit to new data). This includes models trained on wastewater viral load (VL), wastewater detection frequency (DF), and combined DF-VL (ALL) predictor sets. R^2^ was determined for predictions made from January 1, 2022, to April 30, 2022 (prior to testing decline) (a, b), and from May 1, 2022, to December 31, 2022 (after testing decline) (c, d).

**Figure 6.**
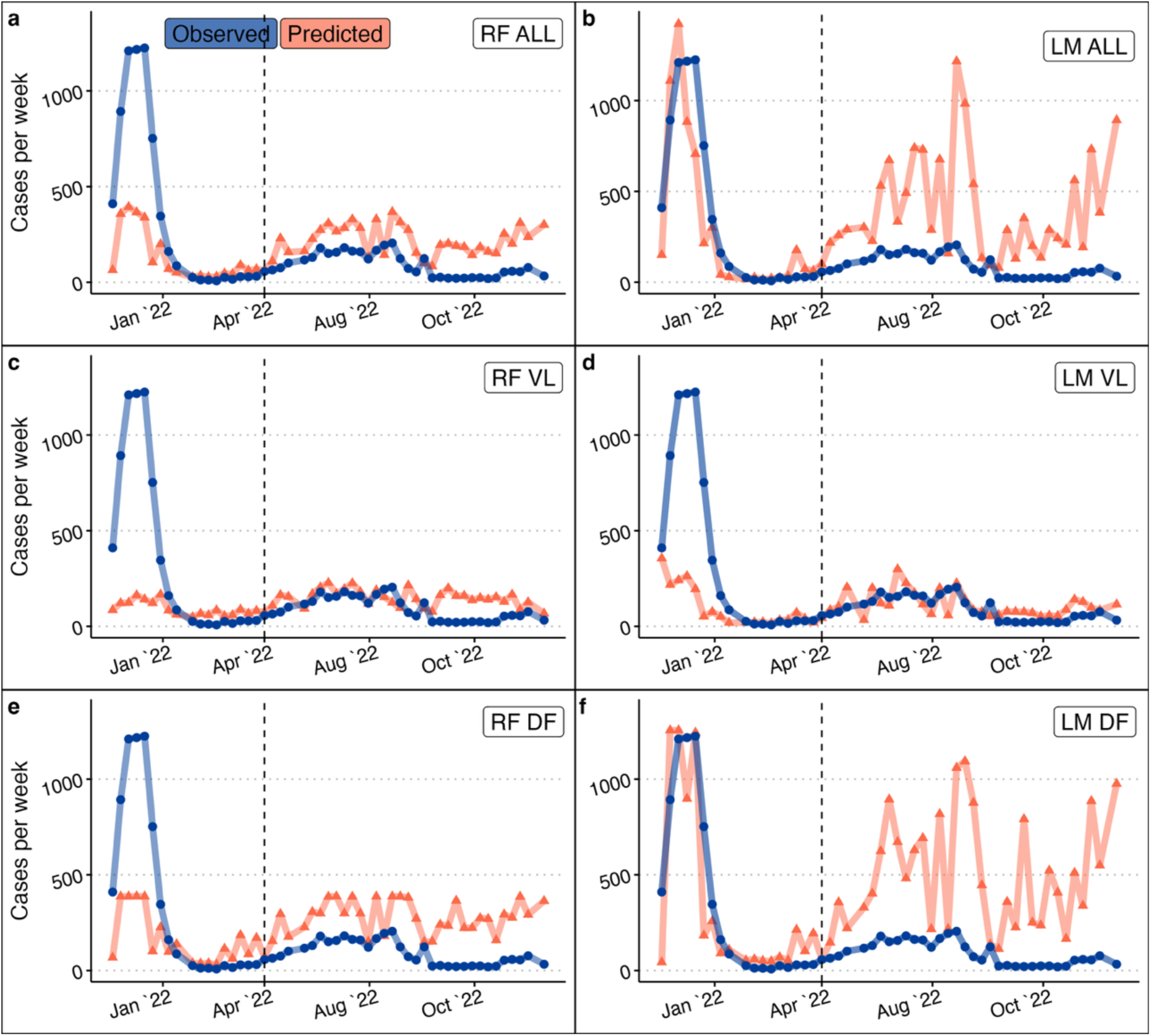
Timeseries of predicted and observed reported cases per week per 100,000 individuals during 2022 (data excluded from model training). Predictions were made using six models: random forest models with ALL (viral load + detection frequency) (a), VL (viral load) (c), and DF (detection frequency) (e) predictor sets, and linear regression models with ALL (b), VL (d), and DF (f) predictor sets. All models were trained on data generated from two extraction replicates. Vertical dotted lines indicate drop in clinical test administration.

Random forest models performed better when fit to training data resamples than linear models (Figure 3; Figure 4; Figure S6; Table S10); however, linear models performed better when making predictions on new data. While random forest models explained just as much variance in new data, linear models did so with much lower prediction error (i.e., lower RMSE) (Figure 6; Table S12). This suggests that linear models may generalize better to new data, despite random forest models showing stronger within-sample performance.

### Predictions after testing decline

Predictions made after April 2022 (after clinical testing efforts diminished) showed weak R^2^ values (0.09 – 0.33) regardless of model type (Figure 5; Figure 6; Table S13). Excluding VL models, predicted case counts after April 2022 were much higher than what was reported. The models that performed best on new data from early 2022 (i.e., LM ALL, LM DF) predicted the highest number of cases in late 2022. For example, the highest number of cases reported per week after early 2022 was 205 per 100,000 (week of August 21), but models predicted 1,216 (LM ALL) and 1,059 (LM DF) cases per 100,000 during this week (Figure 6). These discrepancies suggest that wastewater-based models captured substantial underreporting of COVID-19 cases during periods of reduced clinical testing.

## DISCUSSION

This study employed quantitative PCR methods to monitor SARS-CoV-2 viral load and prevalence in municipal wastewater influent from three neighbouring sewersheds, representing a suburban population of 131,000 over the course of two and a half years. With a dataset, including viral measurements from two gene targets and up to six extraction replicates per sample, we sought to understand trade-offs between simplified versus more complex approaches and ability to predict COVID-19 incidence. We also sought to address certain limitations with quantitative approaches commonly used in wastewater-based epidemiological models.

To do this, we employed data modeling strategies to understand how factors like number of extraction replicates, use of presence/absence (detection frequency) data, and model framework complexity, contributed to prediction accuracy, and strike a balance between prediction accuracy and time to results. Given the current paucity of clinical testing (Rader, 2022; Rubin, 2021) and reporting (CDC, 2025), wastewater testing has become a more consistent surveillance tool (Adams et al., 2024; Naughton et al., 2023). As wastewater increasingly provides the primary data on COVID-19 transmission, actionable predictive models to estimate case rates in a population are needed to better understand disease trends.

### Value of wastewater-based models

When clinical reporting is strong, wastewater signals can robustly track case rates and infection trends within communities, but relationships between wastewater signals and case data deviate when testing and reporting efforts diminish (Boehm et al., 2023; Majumdar et al., 2024). We confirmed in this study that high case reporting in year 1 was likely the critical factor in higher correlations between wastewater variables and reported cases compared to those noted in years 2+, regardless of different detection and quantification limitations. Our models also substantially overpredicted cases in waves 5 and 6 during late 2022.

These findings can be attributed to the overall national shift from PCR testing to at-home rapid antigen testing (Boehm et al., 2023). At-home tests were first introduced in 2021 and became widely available by early 2022, prompting health departments to shift towards vaccination efforts as testing demands decreased (Rader, 2022; Rubin, 2021). Locally, the University of Georgia ended clinical surveillance efforts in May 2022 (Malhotra, 2022), which largely drove testing in the community. As of January 1, 2025, COVID-19 is no longer a nationally notifiable disease in the US, further diminishing the ability to track transmission dynamics through traditional means (CDC, 2025). Wastewater monitoring is a more sustainable and cost-effective approach for tracking infectious diseases in communities (Wolfe, 2022) that is not biased by reporting requirements (CDC, 2025) or healthcare-seeking behaviors (Zheng et al., 2021).

### Considerations for wastewater-based modeling

Rather than concentrating viral particles, which resulted in poor recovery and significant viral loss (Lott et al. 2023), we used a simple direct extraction method from small-volume wastewater influent samples. We performed up to six extraction replicates for each sample (Lott et al., 2023), which allowed us to account for the stochasticity in small volumes and improve detection accuracy (Ahmed et al., 2022; Chen & Bibby, 2023). This study was able to take advantage of the high number of extraction replicates to assess the importance of a range of replicates for predicting COVID-19 cases. We found that two extraction replicates were sufficient for COVID-19 case prediction and while additional replicates did moderately improved predictive models, these additions increased processing costs and time, while not greatly improving prediction accuracy.

Quantitative indicators of SARS-CoV-2 prevalence in wastewater (e.g., viral load, viral concentration, and population normalized viral metrics) have been the primary metrics used in wastewater-based COVID-19 models, to date (Koureas et al., 2021; Li et al., 2023; McManus et al., 2023; Rezaeitavabe et al., 2024). While wastewater viral quantities correlate well with disease incidence (Lott et al., 2023; Medema et al., 2020; Tiwari et al., 2022; Weidhaas et al., 2021), lack of standardized methods (Bivins et al., 2021) and the likelihood of low abundance of viral particles in wastewater between waves (Zhu et al., 2022), can limit the power of quantitative methods. Presence/absence (i.e., detection frequency) of SARS-CoV-2 in wastewater is a simpler measure that also correlates with disease prevalence (Bivins et al., 2022; Lott et al., 2023; Wang et al., 2025) but has been used less frequently.

In the present work, we found that total weekly SARS-CoV-2 detection frequency and average weekly viral load in wastewater exhibited similar relationships to clinically reported COVID-19 cases, regardless of gene target or wastewater catchment. This finding is supported by previous analysis of year 1 data from the same system, which found that wastewater detection frequency measures tended to have similar, if not somewhat stronger, correlations with reported cases compared to viral load (Lott et al., 2023).

Our findings suggest that combining both wastewater viral prevalence measures in predictive models can improve predictive power, compared to sole use of quantifiable measures. Because detection frequency does not rely on precise concentration, this measure is not as sensitive to site-specific variations in wastewater matrices, environmental conditions, and sample handling techniques (Li et al., 2023; Schussman & McLellan, 2022; Weidhaas et al., 2021). Exact concentration of viral RNA in wastewater can be noisy due to low concentrations (Ahmed et al., 2022), inconsistent recovery rates (Ahmed et al., 2022), and variable RT-qPCR (or RT-ddPCR) assay parameters (Bivins et al., 2021) whereas detection frequency can avoid these challenges. Considering that detection frequency is inherently captured by RT-qPCR methods, it would offer a simple approach to add value to models.

SARS-CoV-2 detection frequency may also have value as a sole predictor of clinical incidence. While not as robust as most quantitative-based frameworks, its utility for monitoring SARS-CoV-2 prevalence in wastewater cannot be understated. In this work, we saw that detection frequency models were able to predict clinically reported cases in new data better than solely quantitative-based models. Wastewater detection frequency may be of great value when reliable quantification is not feasible, such as when viral quantities are highly dilute, qPCR or digital droplet PCR technology is not available, or when methodological discrepancies exist, as was the case here. Approaches like endpoint RT-PCR or reverse transcription loop-mediated isothermal amplification (RT-LAMP) offer a promising alternative for detecting viral presence in wastewater that can provide rapid results, forgoing sophisticated quantification equipment. RT-LAMP can also be paired with simple, cost-effective sampling methods (e.g., modified Moore swabs), offering a scalable and practical approach to wastewater monitoring that can indicate clinical incidence comparably to more complex quantitative methods (Bivins et al., 2022).

Previous wastewater-based models have employed a range of algorithms, from simple linear regressions (Koureas et al., 2021) to more complex approaches like ensemble learning (Rezaeitavabe et al., 2025), neural networks (Zamarreño et al., 2024), and time-series models (Jeng et al., 2023). Each modeling framework has distinct strengths depending on the context of use.

However, much of the existing work has prioritized accuracy, with less attention paid to the practical utility of these models to provide early disease warnings or be deployed in low-resource settings.

In our analysis, linear regression models generalized better to new data than random forest models. This may be due in part to the limitations of random forest models when trained on datasets with many zero or low values (Vaughan et al., 2023), as was the case in our study. Linear models, by contrast, offer a practical alternative in such contexts that is computationally efficient, highly interpretable, and suitable for integration into streamlined workflows.

While linear models may not yield the lowest prediction error, making absolute case counts difficult to determine (Koureas et al., 2021), identification of shifts in wastewater trends can provide valuable lead time for preparing public health resources (Vaughan et al., 2023). Their interpretability also enhances transparency and trust, which are often lacking in more opaque “black box” models (James et al., 2021). Although not suitable for all applications, such as forecasting hospitalizations weeks in advance (Li et al., 2023) or identifying critical socioeconomic and demographic factors that influence infection rates (Rezaeitavabe et al., 2025), linear models remain a practical and effective choice in settings where resources are limited, or early warning is the priority.

## CONCLUSIONS

This work explores the impacts of certain SARS-CoV-2 wastewater surveillance strategies on COVID-19 prediction capabilities. Namely, we examined the incorporation of presence/absence measures alongside commonly used quantitative measures of wastewater viral prevalence. We also examined the impact of additional extraction replicates, as well as compared performances between two modeling algorithms. We found that incorporating presence/absence measures in quantitative models could improve predictive power, and that presence/absence measures alone were still able to generalize well to new data. We also determined that an increased number of extraction replicates beyond two improved model performance negligibly, creating additional costs in reagents and processing time while not greatly improving predictions. Finally, we determined that while random forest models exhibited stronger within-sample performances, linear models generalized much better to new data. Together, these findings highlight practical considerations that can inform the development of more efficient, scalable, and accessible wastewater-based monitoring, particularly in resource-limited settings or when early warning is necessary.

## ACKNOWLEDGEMENTS

We appreciate the Athens-Clarke County Public Utilities Department for coordinating the collection of all wastewater samples used in this study.

## AUTHOR CONTRIBUTIONS

L.L., A.H., E.L., and M.L. conceptualized the study and designed the methods. A.F., C.M.D., L.M., M.L., and L.L. performed experiments and documented results. L.L., A.H., and E.L. contributed to data analysis and development of computational models. L.L. wrote the initial draft of the manuscript, and all authors reviewed and edited subsequent versions. E.L. acquired funding and supervised research.

## DATA AVAILABILITY STATEMENT

All data and scripts used in analysis are available on Zenodo (https://doi.org/10.5281/zenodo.15635445).

## ETHICS STATEMENT

None.

