## Supplementary Material for "Balancing Accuracy and Actionability: An Assessment of Minimal- Input Wastewater Models for COVID-19 Prediction"

| Clinical Metric | Mean | Median | SD | Min | Max |
|---|---|---|---|---|---|
| **Reported Cases** | 175.80 | 108.77 | 228.85 | 7.77 | 1224.45 |
| **Tests Administered** | 2146.64 | 1790 | 1505.61 | 414 | 6599 |

**Table S1.** Summary of COVID-19 clinically reported metrics (reported cases per week per 100,000, tests administered per week per 100,000) in Athens-Clarke County during the surveillance period.

| Year | Target | Mean | Median | SD | Min | Max |
|---|---|---|---|---|---|---|
| **Y1** | **N1** | 35.96 | 36.07 | 1.01 | 32.16 | 39.01 |
| | **N2** | 36.01 | 36.08 | 1.01 | 31.23 | 39.03 |
| **Y2+** | **N1** | 36.99 | 37.02 | 1.10 | 31.02 | 39.95 |
| | **N2** | 38.34 | 38.41 | 1.03 | 32.72 | 40.00 |

**Table S2.** Summary of Cq values obtained during the surveillance period for each RT-qPCR SARS-CoV-2 assay.

| Year | Target | # Samples | Samples < LoD | Samples < LoQ |
|---|---|---|---|---|
| **Y1** | **N1** | 916 | 270 (29.5%) | 219 (23.9%) |
| | **N2** | 914 | 272 (29.8%) | 227 (24.8%) |
| **Y2+** | **N1** | 2548 | 2195 (86.1%) | 2195 (86.1%) |
| | **N2** | 2490 | 1758 (70.6%) | 1725 (69.3%) |
| **Total** | | **6868** | **4495 (65.4%)** | **4366 (63.6%)** |

**Table S3.** Summary of RT-qPCR reactions per SARS-CoV-2 assay, indicating total number of reactions (all sample replicates), number of positive detections out of total reactions (at least one technical replicate with Cq < Cq LoD), and number of quantifiable reactions out of total positive detections (at least one technical replicate with Cq < Cq LoQ).

| Year | Target | # Samples | All tech reps < LoD | All tech reps < LoQ |
|---|---|---|---|---|
| **Y1** | **N1** | 916 | 21 (2.3%) | 17 (1.9%) |
| | **N2** | 914 | 15 (1.6%) | 13 (1.4%) |
| **Y2+** | **N1** | 2548 | 1334 (52.4%) | 1331 (52.2%) |
| | **N2** | 2490 | 905 (36.3%) | 875 (35.1%) |
| **Total** | | **6868** | **2275 (33%)** | **2236 (32.6%)** |

**Table S4.** Summary of RT-qPCR reactions per SARS-CoV-2 assay, indicating total number of reactions (all sample replicates), number of reactions where all technical replicates were detectable (Cq < Cq LoD), and number of reactions where all technical replicates were quantifiable (Cq < Cq LoQ).

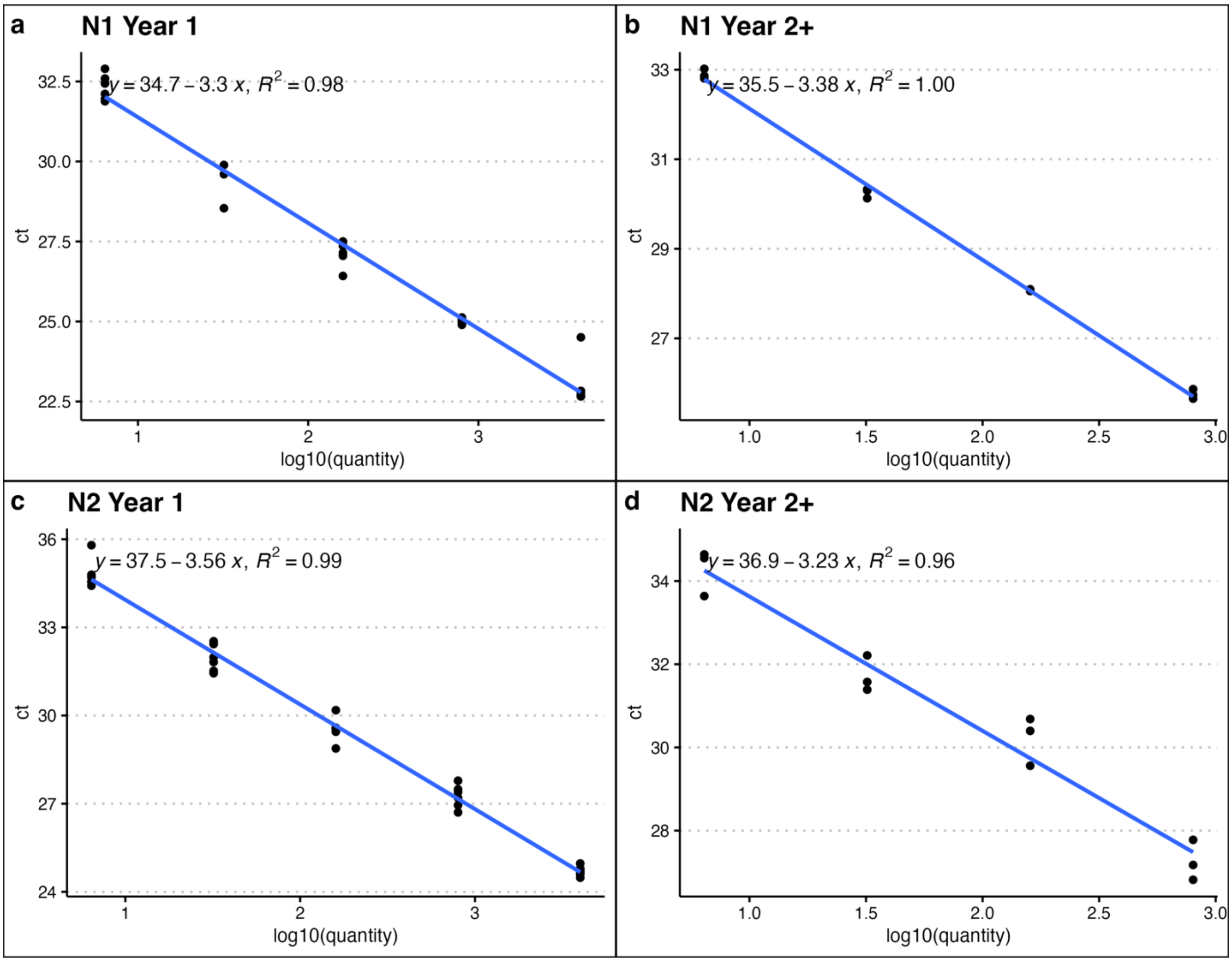


**Figure S1.** Standard curves for year 1 (StepOne) and year 2+ (CFX) instrumentation and each gene target (N1, N2).

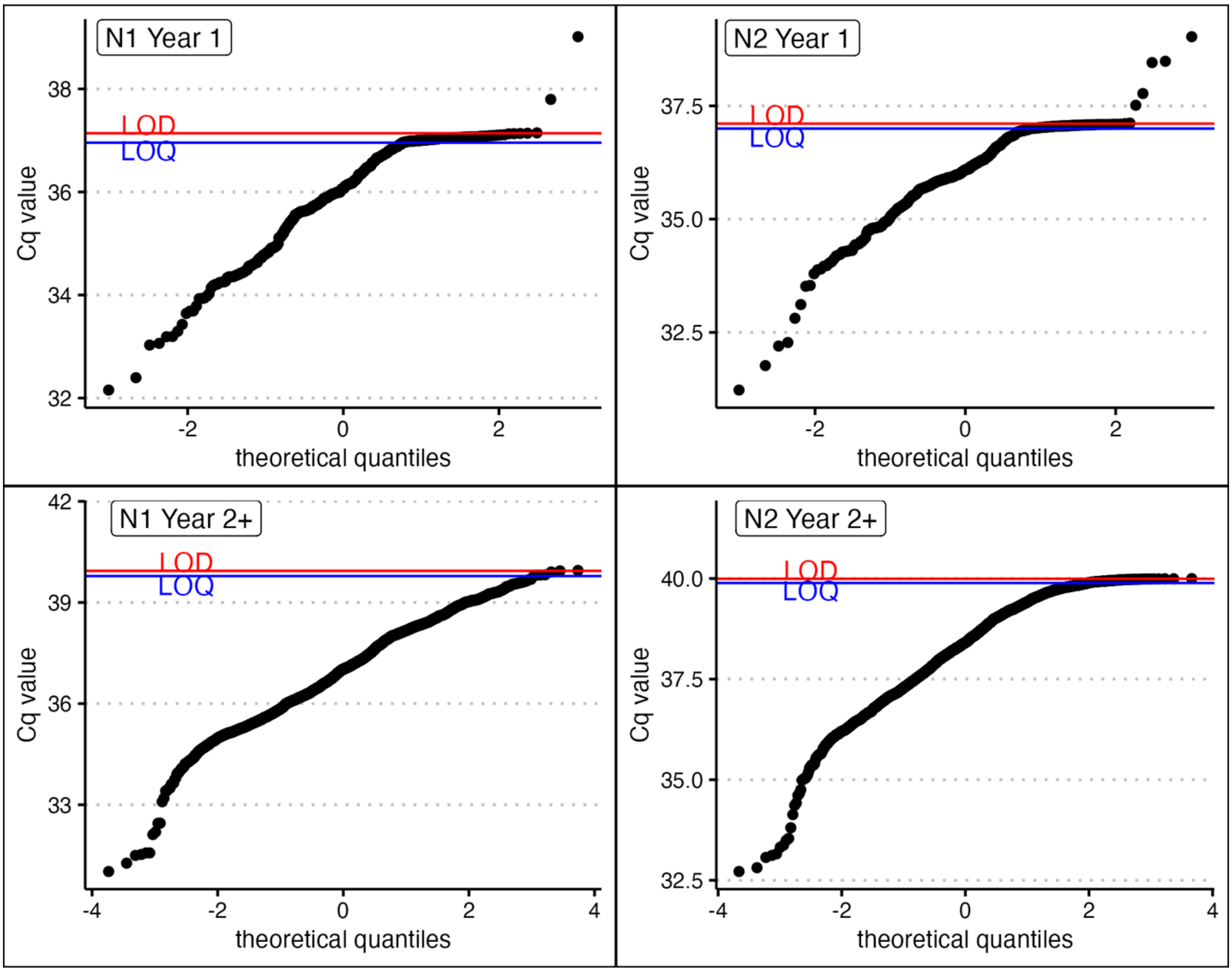


**Figure S2.** Normal Q-Q plot of Cq values for year 1 (StepOne) and year 2+ (CFX) instrumentation and each gene target (N1, N2).

| Year | Target | Cq | Copies/L |
|---|---|---|---|
| **Y1** | **N1** | 37.14 | 3.23E+6 |
| | **N2** | 37.11 | 2.30E+7 |
| **Y2+** | **N1** | 39.94 | 4.19E+4 |
| | **N2** | 39.99 | 9.15E+4 |

**Table S5.** Limit of detection (LoD) for each RT-qPCR SARS-CoV-2 assay.

| Year | Target | Cq | Copies/L |
|---|---|---|---|
| **Y1+** | **N1** | 36.96 | 3.67E+6 |
| | **N2** | 37.00 | 2.47E+7 |
| **Y2+** | **N1** | 39.78 | 4.65E+4 |
| | **N2** | 39.89 | 9.87E+4 |

**Table S6.** Limit of quantification (LoQ) for each RT-qPCR SARS-CoV-2 assay.

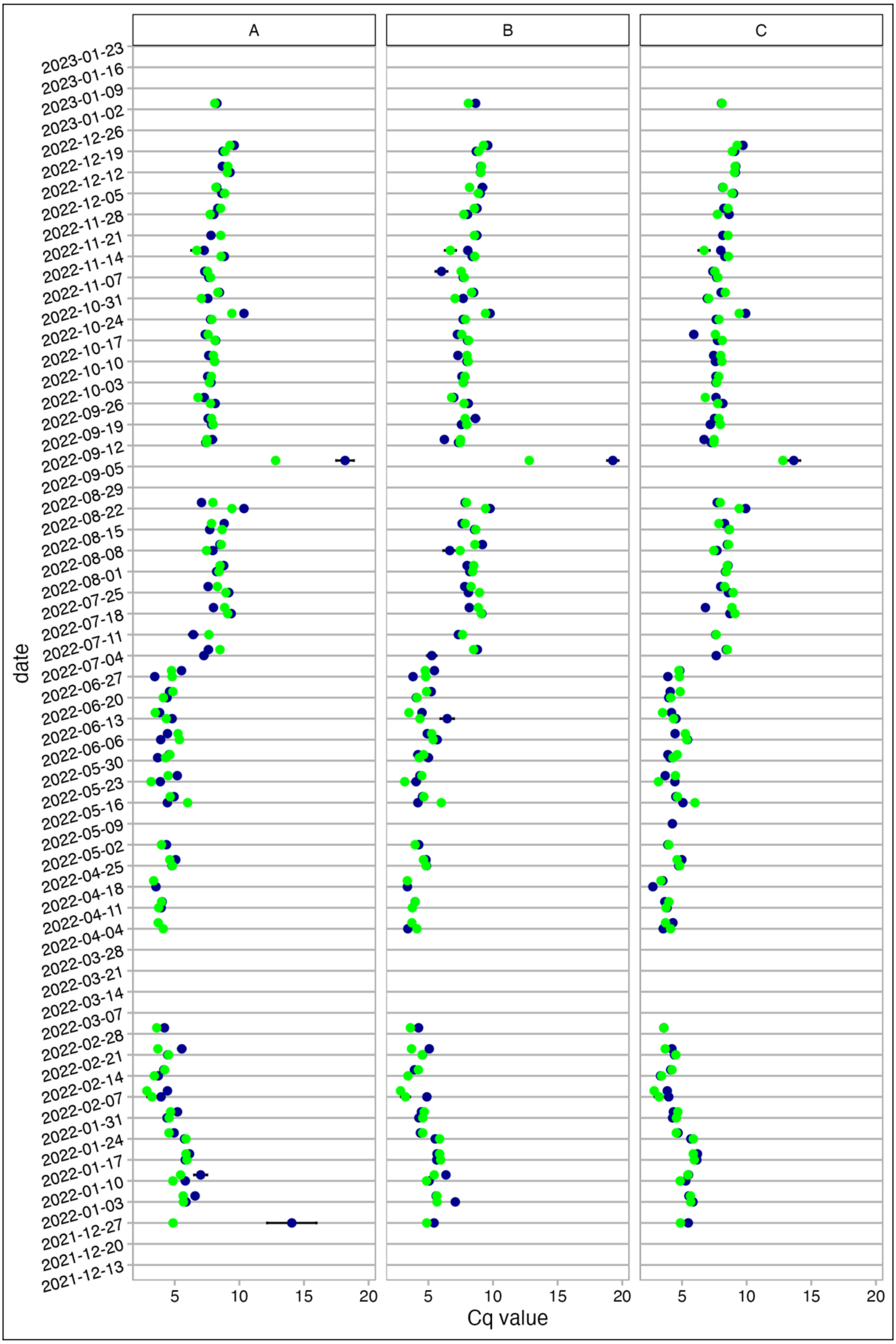


**Figure S3.** Assessment of inhibition in RNA extracts from wastewater samples in year 2+, organized by collection date and water reclamation facility. Average Cq values shown represent BCoV spiked into wastewater samples (blue) and controls (green), plus error bars to indicate standard deviation of Cq values. See Lott et al. (2023) for assessment of inhibition in year 1 extracts (Lott et al., 2023)

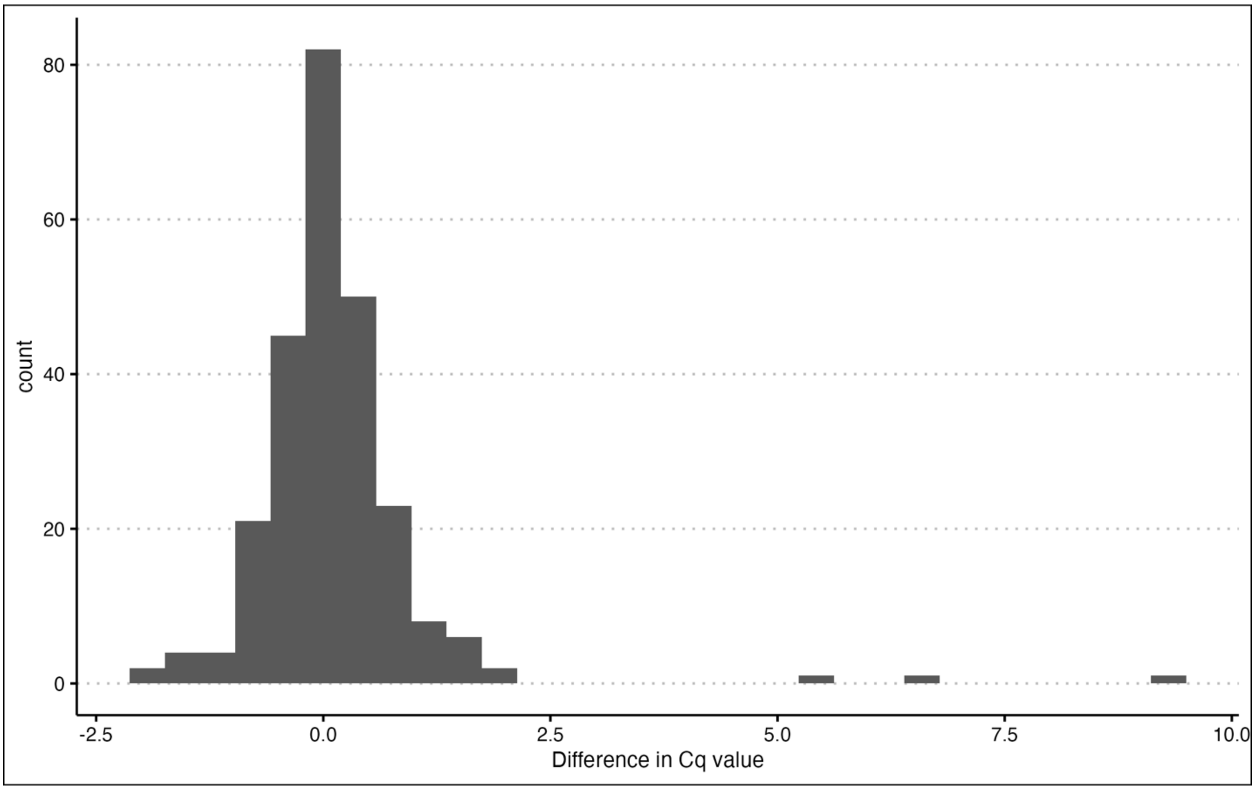


**Figure S4.** Assessment of inhibition in RNA extracts from wastewater samples in year 2+, shown as the distribution of differences between sample Cq values and BCoV control Cq values. See Lott et al. (2023) for assessment of inhibition in year 1 extracts (Lott et al., 2023).

| WRF | Target | Mean | Median | SD | Min | Max |
|---|---|---|---|---|---|---|
| A | **N1** | 9.09E+12 | 6.17E+12 | 1.38E+13 | 4.40E+11 | 1.04E+14 |
| | **N2** | 2.88E+13 | 3.52E+12 | 9.15E+13 | 7.33E+11 | 9.37E+14 |
| **B** | **N1** | 7.11E+12 | 5.34E+12 | 9.33E+12 | 2.96E+11 | 7.69E+13 |
| | **N2** | 1.92E+13 | 6.29E+12 | 4.86E+13 | 5.02E+11 | 4.82E+14 |
| **C** | **N1** | 3.73E+12 | 2.51E+12 | 1.18E+13 | 1.85E+11 | 1.33E+14 |
| | **N2** | 6.74E+12 | 1.95E+12 | 1.33E+13 | 2.99E+11 | 9.32E+13 |

**Table S7.** Summary of SARS-CoV-2 wastewater viral load per wastewater reclamation facility and gene target per week.

| WRF | Target | Mean | Median | SD | Min | Max |
|---|---|---|---|---|---|---|
| **A** | **N1** | 0.44 | 0.39 | 0.35 | 0 | 1 |
| | **N2** | 0.33 | 0.28 | 0.29 | 0 | 1 |
| **B** | **N1** | 0.53 | 0.50 | 0.38 | 0 | 1 |
| | **N2** | 0.44 | 0.33 | 0.36 | 0 | 1 |
| **C** | **N1** | 0.51 | 0.47 | 0.37 | 0 | 1 |
| | **N2** | 0.43 | 0.38 | 0.34 | 0 | 1 |

**Table S8.** Summary of SARS-CoV-2 wastewater detection frequency per wastewater reclamation facility and gene target per week.

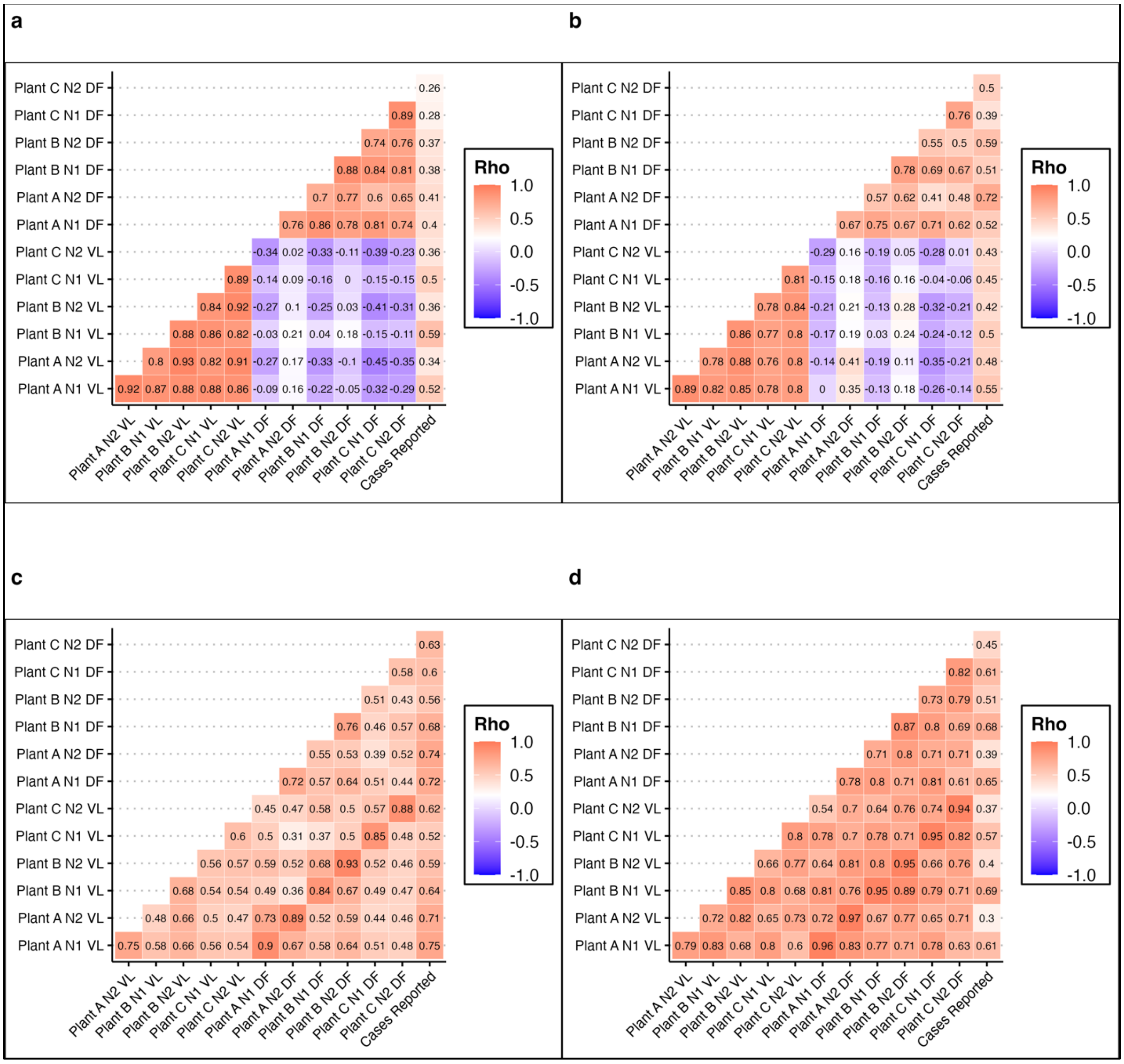


**Figure S5.** Matrix of Spearman correlation coefficients between all SARS-CoV-2 wastewater measures and COVID-19 reported cases (per week) in the full dataset (a), data used for model training (b), data from year 1 (c), and data from year 2+ (d).

| Feature | trees | mtry | min_n |
|---|---|---|---|
| **Detection freq.** | 1000 | 6 | 19 |
| **Viral load** | 1000 | 1 | 6 |
| **Combined** | 1000 | 5 | 3 |

**Table S9.** Random forest model hyperparameters for each feature selection. Specifically, the number of trees, the number of predictors sampled at each split (mtry), and the number of data points required for further splitting of a node (min_n).

| Model type | Predictor set | Extraction rep. | RMSE | SE | $R^2$ | SE |
|---|---|---|---|---|---|---|
| **Linear regression** | **Detection freq.** | **1** | 147.68 | 1.649 | 0.28 | 0.009 |
| | | **2** | 155.06 | 1.955 | 0.38 | 0.009 |
| | | **3** | 166.27 | 3.170 | 0.39 | 0.007 |
| | | **4** | 178.09 | 3.646 | 0.40 | 0.008 |
| | | **5** | 162.99 | 2.717 | 0.43 | 0.007 |
| | | **6** | 164.24 | 2.743 | 0.42 | 0.007 |
| | **Viral load** | **1** | 155.62 | 2.306 | 0.18 | 0.007 |
| | | **2** | 137.11 | 2.292 | 0.43 | 0.006 |
| | | **3** | 140.62 | 2.478 | 0.36 | 0.006 |
| | | **4** | 140.00 | 2.401 | 0.37 | 0.005 |
| | | **5** | 140.52 | 2.397 | 0.37 | 0.006 |
| | | **6** | 139.69 | 2.341 | 0.38 | 0.005 |
| | **Combined** | **1** | 153.78 | 3.811 | 0.44 | 0.013 |
| | | **2** | 143.08 | 4.116 | 0.60 | 0.008 |
| | | **3** | 120.28 | 2.747 | 0.70 | 0.005 |
| | | **4** | 128.49 | 3.253 | 0.69 | 0.005 |
| | | **5** | 125.00 | 3.445 | 0.69 | 0.006 |
| | | **6** | 129.74 | 3.838 | 0.69 | 0.006 |
| **Random forest** | **Detection freq.** | **1** | 146.74 | 1.737 | 0.26 | 0.012 |
| | | **2** | 125.42 | 1.573 | 0.46 | 0.007 |
| | | **3** | 125.05 | 1.638 | 0.45 | 0.007 |
| | | **4** | 127.90 | 1.939 | 0.42 | 0.006 |
| | | **5** | 121.42 | 1.722 | 0.48 | 0.005 |
| | | **6** | 121.54 | 1.729 | 0.48 | 0.006 |
| | **Viral load** | **1** | 136.16 | 2.130 | 0.42 | 0.015 |
| | | **2** | 118.61 | 2.119 | 0.61 | 0.007 |
| | | **3** | 115.47 | 2.050 | 0.61 | 0.007 |
| | | **4** | 117.67 | 2.119 | 0.59 | 0.007 |
| | | **5** | 118.38 | 2.112 | 0.58 | 0.007 |
| | | **6** | 116.40 | 2.130 | 0.60 | 0.007 |
| | **Combined** | **1** | 124.56 | 2.014 | 0.51 | 0.014 |
| | | **2** | 103.67 | 1.859 | 0.67 | 0.006 |
| | | **3** | 98.90 | 1.871 | 0.69 | 0.005 |
| | | **4** | 99.78 | 2.007 | 0.69 | 0.005 |
| | | **5** | 98.06 | 1.915 | 0.70 | 0.005 |
| | | **6** | 97.07 | 1.869 | 0.71 | 0.005 |
| **Null Model** | **Combined** | **6** | 175.12 | 2.609 | 0.07 | 0.003 |

**Table S10.** Mean cross-validated model performance metrics and standard error of each model when resamples of the training data.

| Model type | Predictor set | RMSE | SE | $R^2$ | SE |
|---|---|---|---|---|---|
| **Linear regression** | **Detection freq.** | 162.39 | 1.725 | 0.38 | 0.009 |
| | **Viral load** | 142.26 | 1.112 | 0.35 | 0.014 |
| | **Combined** | 133.4 | 2.094 | 0.64 | 0.017 |
| **Random forest** | **Detection freq.** | 128.01 | 1.584 | 0.43 | 0.014 |
| | **Viral load** | 120.45 | 1.298 | 0.57 | 0.012 |
| | **Combined** | 103.68 | 1.747 | 0.66 | 0.013 |

**Table S11.** Cross-validated model performance metrics and standard error averaged by predictor variable-model type combination (training data).

| Model type | Predictor set | Extraction rep. | RMSE | $R^2$ |
|---|---|---|---|---|
| **Linear regression** | **Detection freq.** | **2** | 208.36 | 0.8 |
| | **Viral load** | **2** | 468.23 | 0.57 |
| | **Combined** | **2** | 220.3 | 0.79 |
| **Random forest** | **Detection freq.** | **2** | 404.26 | 0.74 |
| | **Viral load** | **2** | 506.54 | 0.65 |
| | **Combined** | **2** | 410.90 | 0.86 |

**Table S12.** Model performance metrics and standard error of each model when fit to new data (January 2022 – April 2022).

| Model type | Feature | Biological rep. | RMSE | $R^2$ |
|---|---|---|---|---|
| **Linear regression** | **Detection freq.** | **2** | 487.89 | 0.11 |
| | **Viral load** | **2** | 61.1 | 0.33 |
| | **Combined** | **2** | 410.2 | 0.21 |
| **Random Forest** | **Detection freq.** | **2** | 204.76 | 0.09 |
| | **Viral load** | **2** | 83.85 | 0.11 |
| | **Combined** | **2** | 143.72 | 0.23 |

**Table S13.** Model performance metrics and standard error of each model when fit to data after clinical test administration declined (May 2022 – January 2023).

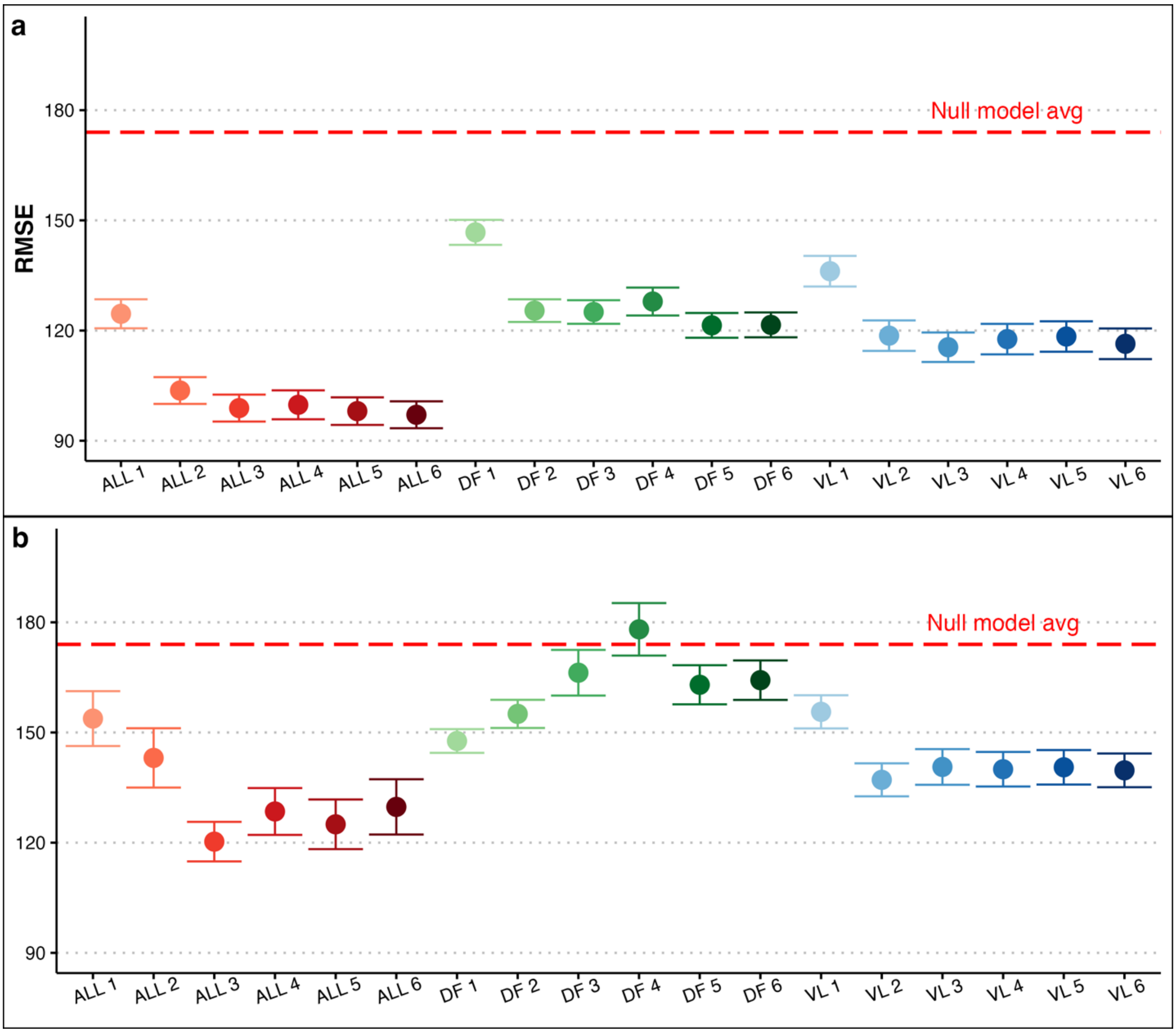


**Figure S6.** Average cross-validated RMSE and 95% CI for (a) all random forest models and (b) all linear regression models. This includes models trained on wastewater viral load (VL), wastewater detection frequency (DF), and combined DF-VL (ALL), as well as randomly selected extraction replicates (1-6).

| EMMI Reporting Element | Location |
| --- | --- |
| **ENVIRONMENTAL SAMPLING** | |
| Sampling procedure, sample amount and number | Methods |
| Sampling locations and dates | Methods |
| Sample storage conditions and duration | Methods |
| Sampling negative control* | Methods |
| Sampling positive control* | Methods |
| **SAMPLE TREATMENT** | |
| Treatment procedure and reagents | Methods |
| Treatment negative control* (e.g. filter eluent) | Methods |
| Treatment positive control* (may be combined with sampling positive control) | Methods |
| **SAMPLE REDUCTION** | |
| Reduction procedure | NA |
| Concentration factor | NA |
| Reduction negative control* (may be combined with treatment negative control) | NA |
| Reduction positive control* (may be combined with sampling and treatment positive control) | NA |
| **NUCLEIC ACID EXTRACTION** | |
| Extraction procedure | Methods |
| Concentration factor: amount extracted, and amount obtained | NA |
| Extract storage conditions and duration | Methods |
| Extraction negative control | Methods |
| Extraction positive control | Methods |
| **REVERSE TRANSCRIPTION** | |
| One or two step reaction | Methods |
| RT reaction temperatures and times | Methods |
| RT reaction reagents and concentrations | Methods |
| Priming method | Methods |
| Reaction volume, template amount added | Methods |
| cDNA storage conditions and duration | NA |
| RT negative control | Methods |
| RT positive control | Methods |
| Inhibition assessment procedure | Methods |
| Number of samples tested and found inhibited | Supplemental Figure S3, S4 |
| Inhibition control description (if control used) | Methods |
| **qPCR** | |
| Target gene name, amplicon length | Methods |

| | |
|---|---|
| Thermocycling temperatures and times | Methods |
| Master mix composition, vendors, concentrations | Methods |
| Additives, vendors, and concentrations | Methods |
| Template amount added, pre-treatment (if any) | Methods |
| Primer conc., vendor, sequence, reference | Methods |
| Hydrolysis probe concentration, dye and quencher, vendor, sequence, reference | Methods |
| Instrumentation | Methods |
| Amplicon confirmation method (e.g. probe, melt curve, gel, sequencing) | NA |
| Equivalent volume of sample analyzed by PCR | Results |
| qPCR negative control | Methods |
| qPCR positive control | Methods |
| Inhibition assessment procedure | Methods |
| Number of samples tested and found inhibited | Supplemental Figure S3, S4 |
| Inhibition control description (if control used) | Methods |
| **ANALYSIS - qPCR** | |
| Accounted for negative controls that failed (e.g. Cq value separation between positives and sample contamination) | NA |
| Technical replicates: number, calculations, and summary statistics performed | Methods, Supplemental Table S2, S3, & S4 |
| Calibration standards, description and source | Methods |
| Standards quantification method | Methods |
| Calibration curve slope (i.e., PCR efficiency) | Supplemental Figure S1 |
| Calibration curve R^2 | Supplemental Figure S1 |
| Lowest standard measured of 95% LOD | NA |
| Cq value determination method (auto or manual threshold placement) | Methods |

**Table S14.** Environmental Microbiology Minimum Information (EMMI) Guidelines (Borchardt et al., 2021).
